# Genetic Determinants of Major Depressive Disorder among Adult Persons Living With HIV in Uganda

**DOI:** 10.1101/2025.03.19.25324246

**Authors:** Olga Nsangi Tendo, Ronald Galiwango, Eugene Kinyanda, Martha Sajatovic, Mark Kaddumukasa, Martin Kaddumukasa, Elly Katabira, Catherine Nabbumba, Soraya Seedat, Sian M.J. Hemmings, Segun Fatumo, Allan Kalungi

## Abstract

**Background:** Major Depressive Disorder is complex and moderately heritable, with heritability estimates ranging from 30% to 40%. Major depressive disorder (MDD) is a significant comorbidity in people living with HIV (PLWHIV), where it is associated with non-adherence to anti-retroviral treatment, increased risk of suicide-related behaviors among others. This study investigated the genetic risk loci associated with MDD among adult PLWHIV in Uganda.

**Methods:** The case-control study analyzed 282 samples (139 MDD cases and 143 controls), who were assessed for MDD using the Mini International Neuropsychiatric Interview. The DNA of study participants was genotyped at the Center for Proteomic and Genomic Research in South Africa using the H3Africa array (version2). Data were analyzed using PLINK2 and GEMMA softwares for quality control and genome-wide association analysis respectively, followed by functional mapping with FUMA software.

**Results:** While no significant single nucleotide polymorphisms (SNPs) were identified at the genome-wide threshold, three independent SNPs were found to be suggestively associated with MDD with the most significant being rs4375916 on chromosome 2 (p=1.086×10^−7^), followed by rs1075618 on chromosome 3 (p=3.093×10^−7^) and rs953855 on chromosome 13 (p=4.906×10^−7^). These SNPs have been found to be linkage disequilibrium (LD) with SNPs previously associated with other psychiatric conditions like Alzheimer’s, alcohol use disorder, and bipolar disorder, although they have not previously been linked to MDD. Gene-set enrichment analysis revealed associations with key biological processes including synaptic organization suggesting potential involvement in the pathophysiology of MDD.

**Conclusion:** This study provides preliminary evidence for genetic contributors to MDD in PLWHIV of African ancestry. While larger, adequately powered studies are needed, our findings emphasize the importance of investigating psychiatric genetics in HIV populations, where depression represents a critical comorbidity influencing treatment outcomes.

## Background

Major depressive disorder (MDD) is a common, disabling neuropsychiatric disorder, with multifactorial etiology [1]. The disorder is responsible for 50 million years lived with disability (YLD) worldwide, accounting for 7.5% of global total YLD, and is therefore regarded as the largest contributor to non-fatal health loss [2]. Family and twin studies indicate a strong genetic contribution to the etiology of MDD, with twin-based heritability estimates ranging from 30% to 40% [3].

Major depressive disorder is a common cause of psychiatric morbidity in persons living with human immunodeficiency virus (PLWHIV) [4], [5], with 39% of human immunodeficiency virus (HIV) patients having MDD [6]. In 2019, there were 25.6 million PLWHIV in sub-Saharan Africa (SSA), with a prevalence of MDD ranging from 9% to 32%, in this population [7] . MDD has been associated with negative behavioral and clinical outcomes in HIV-affected individuals, including rapid HIV disease progression and mortality, poor adherence to antiretroviral therapy, risky sexual behavior, and decreased utilization of health facilities [8], [9]. Despite the impact of MDD on PLWHIV, it has been largely overlooked as a priority within integrated HIV care services in sub-Saharan Africa [5]. Previous studies have shown that depression is not only associated with higher HIV viral loads and lower CD4 cell counts [10] but also hastens the progression to AIDS and elevates the risk of mortality [10]. Furthermore, MDD has been reported to reduce adherence to antiretroviral therapy (ART), weaken its therapeutic effects, and compromise the medication outcomes at both individual and population levels [7], [10]. Adherence to ART medications is instrumental in treatment effectiveness and clinical outcomes, while ART interruption and discontinuation worsen physical functioning, and along with depressive behaviors, this could result in further impairments in social relationships and a consequential reduced overall quality of life [10]. The human immunodeficiency virus/acquired immunodeficiency syndrome and MDD exhibit overlapping symptoms such as fatigue, sleep problems, bereavement, loss of appetite and cognitive impairment [10] and this has led to the under diagnosis of MDD in HIV and in many instances, HIV too goes undiagnosed. This misdiagnosis of MDD in PLWHIV has been under-recognized [10] and is often erroneously attributed to psychological distress related to the HIV diagnosis.

Several studies have investigated socio-demographic, psychosocial and clinical risk factors of MDD in HIV but there remains a critical gap in understanding its genetics which is crucial for unraveling the biological mechanisms underlying the disorder [11]. This has been addressed through this genome-wide association study (GWAS) to identify genetic loci associated with MDD among the study participants.

Genome-wide approaches are now widely used in medical research; however, few such studies have been conducted in low- and middle-income countries (LMICs) [12]. Recent GWAS meta-analyses have identified common genetic variants that are associated with MDD [3], [13], [14], [15], however, most of these have been conducted among individuals of European ancestry, and those that included Africans [14] primarily involved African-admixed populations in America and Europe, who may not accurately reflect the genetic diversity of continental Africans [16], [17]. These populations often have genetic contributions from Europeans, Native Americans, or other groups, which can skew results in studies aimed at understanding genetic diversity or disease susceptibility specifically in populations of purely African descent [18]. This admixture can obscure signals of genetic variants that are more prevalent or functionally important in continental African populations, leading to inaccurate or incomplete conclusions about African genetics [19]. There is a need to understand the genetics of depression not only in European populations, but also in other ethnic groups since several studies have shown that psychiatric disorders may vary among different ethnic groups [20] calling for more research on the etiology of MDD in understudied populations, especially Africans [21]. African populations harbor the greatest genetic diversity globally [22], making them a rich resource for identifying rare and common genetic variants that may not exist in European or other populations [22]. There is therefore a need for studies on the genetic factors that underlie MDD among populations of African ancestry [23] as they may facilitate the identification of novel genetic markers that could represent novel mechanistic pathways that could enable the development of therapeutics that are more efficacious and have minimal side effects [24]. Additionally, findings from such studies can be shared with larger study consortia such as the Psychiatric Genomics Consortium to contribute to wider research efforts towards understanding the genetic risk for MDD [24].

## Materials and methods

### Study design

A total of 282, (139 cases and 143 controls) samples used for the GWAS were selected from samples of a genetics sub-study [25] of the EDCTP-funded project (grant number: Ta. K. 40,200.01) as shown in figure1. The EDCTP-funded study focused on mental health among HIV-positive patients at TASO clinics in Entebbe and Masaka, Uganda

### Identifying samples for the genetics sub-study and their description

The flow chart in **figure 1** below summarizes how the samples used for the GWAS study were identified

**Figure 1:**
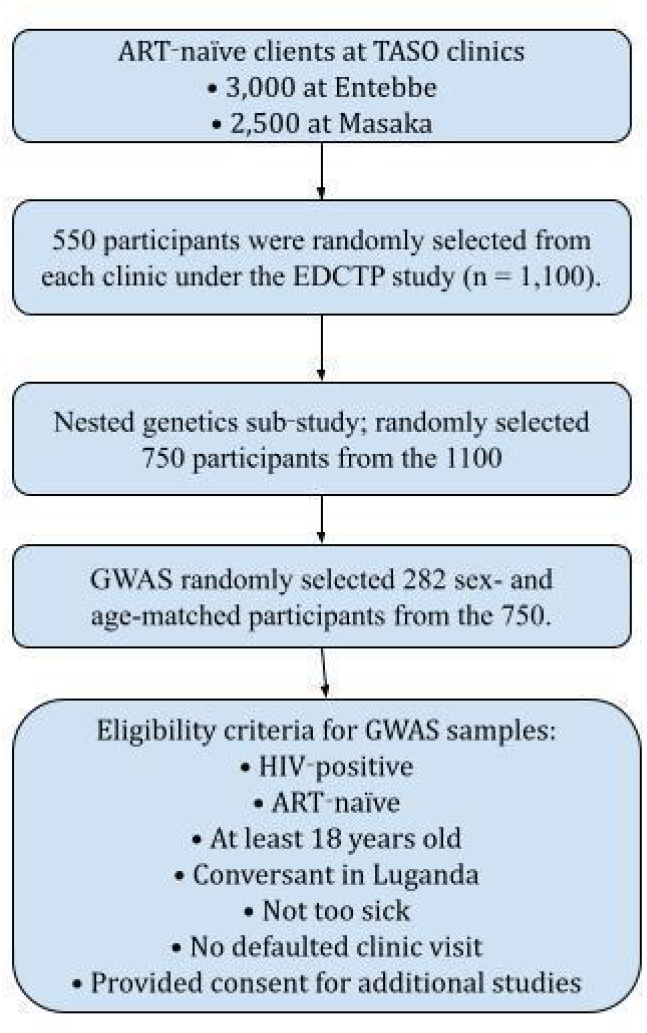
Sample selection flow for GWAS on MDD among HIV-positive clients at TASO Entebbe and Masaka. From 5,500 clients, 1,100 were enrolled in the EDCTP study; 750 entered a nested genetics sub-study, and 282 participants, (141 cases and 141 controls) were included in the GWAS

All participants described in figure 1 had been assessed for MDD at baseline, month 6, and month 12. Assessment for MDD was conducted by psychiatric nurses using the MDD module of the Mini International Neuropsychiatric Interview-Plus (M.I.N.I. Plus) [26], a structured tool for diagnosing mental disorders according to DSM-IV.

### DNA extraction

The DNA was extracted from 3 mL of whole blood samples using the Qiamp Mini DNA Extraction Kit (Qiagen, Germany), following manufacturer’s instructions and suspended in 50 µL of nuclease-free water to prevent degradation. The quality and quantity of the DNA were assessed using spectrophotometer absorbance ratios (260:280) and stored at -20°C until shipment. The samples were then shipped on dry ice to the Center for Proteomics and Genomic Research (CPGR) in South Africa for genotyping. Genotyping was performed using the Infinium LCG Assay protocol with the custom H3Africa v2 array BeadChip (Illumina product code: ILL-15056944), covering over two million SNPs across various African population groups.

### Data management and Statistical analysis

#### Raw data quality control

The raw data in intensity data (IDAT) format was assessed using GenomeStudio 2.0 software [27]. To load the IDAT files, a sample sheet (containing sample information and indexes) and a manifest file (describing SNPs or probes on the BeadChip) were required. Samples with call rates of 0.95 or higher passed the quality control (QC). The GenTrain score, which measures the quality of SNP calling (ranging from 0.0 to 1.0), was also used for data quality assessment, with reliable scores between 0.5 and 1.0. Only SNPs that met these criteria were retained for further analysis. The IDAT files were shared with the Mental Health Unit at MRC/UVRI Uganda and Makerere University for genome-wide association study (GWAS) analysis, where they were processed using GenomeStudio to generate PLINK-compatible file formats (.bed, .bim, .fam) containing genotype, SNP, and participant information.

### Quality control and cleaning of transformed data

Data cleaning and QC involved assessing the transformed genetic data for missingness, duplicates, and potential errors such as sample contamination or population stratification. This step ensures that only high-quality data are included in subsequent analyses [13], [28]. The genotype data was filtered at both the SNP and sample levels using criteria such as SNP-level missingness, individual-level missingness, minor allele frequency (MAF), Hardy-Weinberg equilibrium (HWE), SNP heterozygosity, relatedness, and population stratification using Plink 1.9 software [29]. The thresholds for QC are shown in **Figure 2** in the results section and these were selected with guidance from [13].

**Figure 2:**
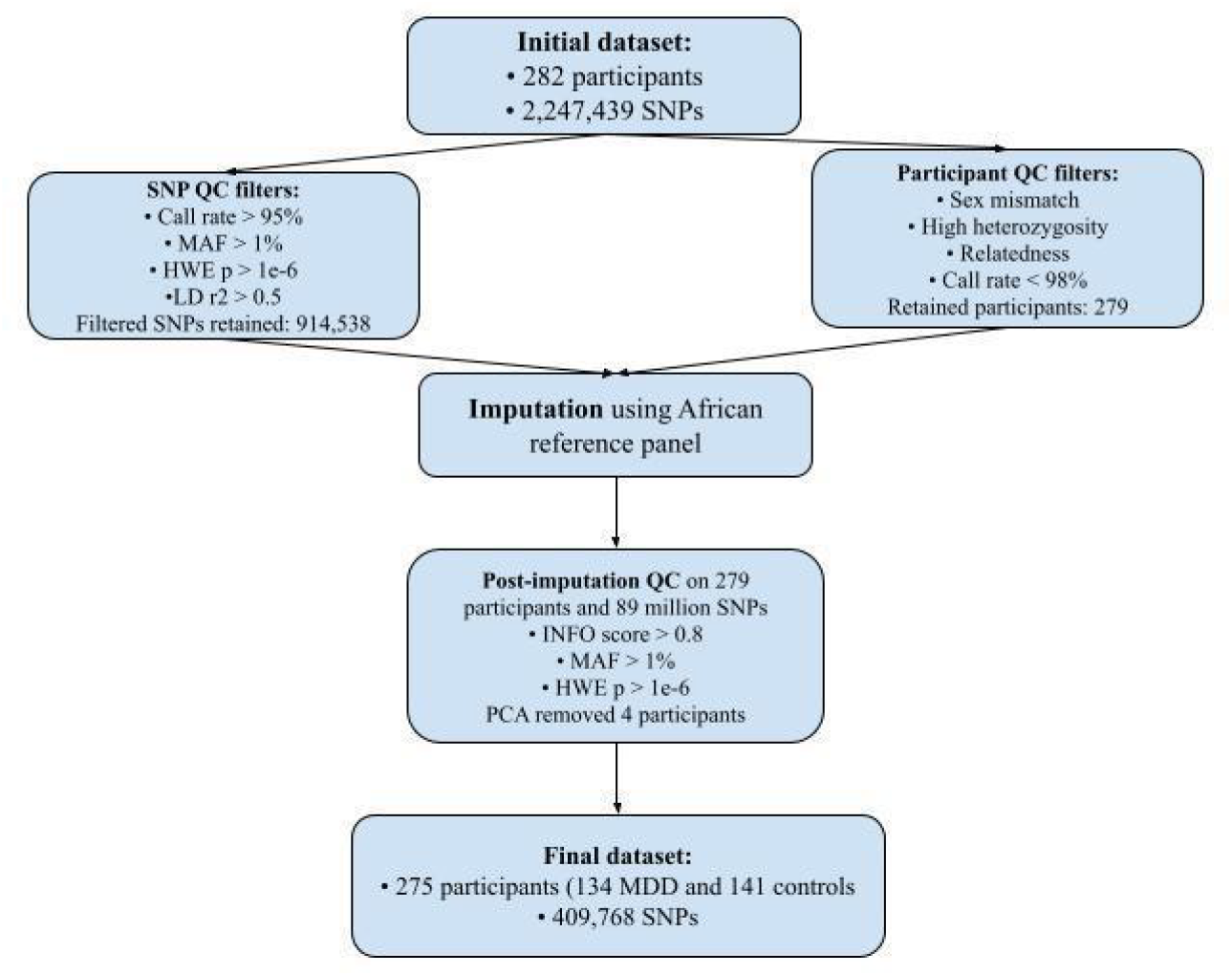
Quality control steps done on the genotype data for 282(141 cases and 141 controls) HIV-positive clients at TASO Entebbe and Masaka before running a genome wide analysis.

To assess population structure and ensure sample quality, we performed principal component analysis (PCA) using PLINK. PCA was first conducted on our cohort alone to identify population structure and potential outliers. We then repeated the PCA including samples from the 1000 Genomes Project (Phase 3) (https://www.internationalgenome.org/data-portal/data-collection/phase-3) to evaluate the ancestry composition of our cohort in a global context. The combined dataset included samples from our cohort, African, European, and other continental populations as defined in the 1000 Genomes reference panel. Genotypes were pruned for linkage disequilibrium prior to analysis. Additionally, also the genomic inflation factor (λGC) was calculated to assess potential residual population stratification. After data cleaning and QC, missing and un-typed SNPs were imputed using reference data from the African Genome Resource via the Sanger Imputation Server [30], ensuring accurate imputation of population-specific variants. Imputed SNPs were also subjected to post imputation QC filtering for MAF ≥ 0.01, linkage disequilibrium (LD, r^2^ ≥ 0.5), and removing SNPs with low imputation accuracy (info score < 0.7). The imputation process ensured comprehensive genome coverage, allowing for the inclusion of variants that may be specific to African populations otherwise absent from the genotyping array.

### Genome-wide association analysis

The GWAS examined the relationship between each SNP and MDD, leveraging a linear model to identify loci associated with the disorder [31]. The GWAS was performed using GEMMA software [31], with MDD. This threshold was based on the fact that it is a less stringent cutoff which balances exploratory power with statistical rigor, highlighting loci that might merit further investigation in replication studies or follow-up which applied a univariate linear mixed model to test associations between SNPs (both genotyped and imputed) and MDD. The model adjusted for age, sex, and principal components. The genome-wide significance threshold was set at (p ≤ 5×10^−8^). A suggestive threshold (p ≤ 1×10^−6^) was explored to identify SNPs that exhibited a trend towards significant association research [32].

### Post-analytic visualization

Post-analytic visualization included a Manhattan plot to display significant SNPs by chromosomal position, a quantile-quantile (Q-Q) plot to assess deviations between observed and expected p-value distributions as shown in **Figures 4 and 5** respectively. A PCA plot for this cohort is included in supplementary data as figure S1.png and two PCA plots for this cohort overlaid on the 1000 genome participants is shown in **Figure 3**. These visuals were generated in R software using the ggplot2 and qqman packages [33], [34], [35]. Scripts to plot the visuals include supplementary data as PCAs.R and 1KG_Uganda_Merged_PCA.R.

**Figure 3:**
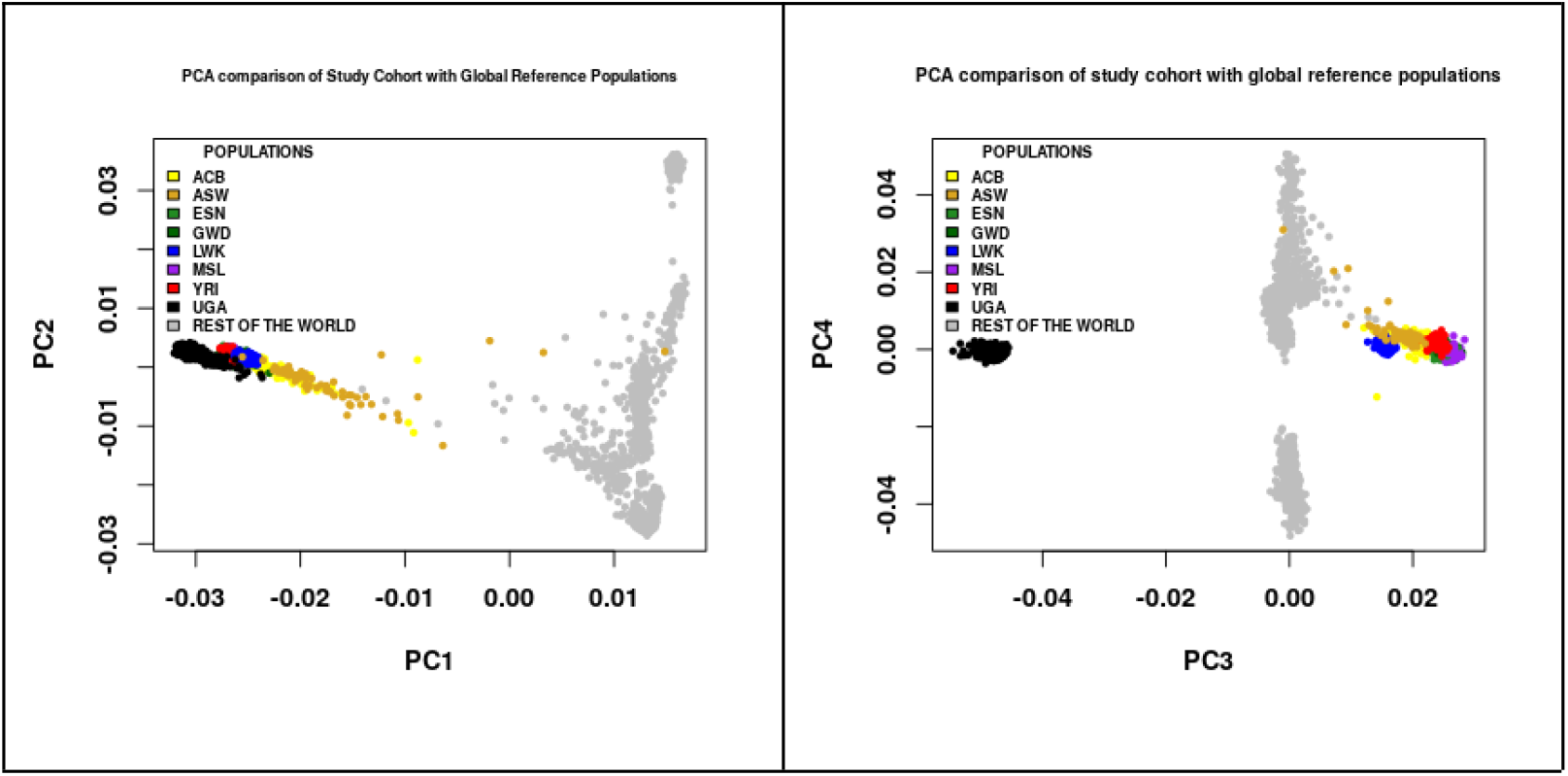
PCA of the study cohort with global reference populations. Left panel: PC1 vs PC2 shows major continental-level genetic structure. Right panel: PC3 vs PC4 highlights finer-scale population substructure, particularly within African groups. Each color represents a different African population ACB - African Caribbean in Barbados, ASW - African Ancestry in Southwest USA, ESN - Esan in Nigeria, GWD - Gambian in Western Divisions, The Gambia, LWK - Luhya in Webuye, Kenya, MSL - Mende in Sierra Leone, YRI - Yoruba in Ibadan, Nigeria, UGA - Uganda (the study cohort); grey dots indicate individuals from reference populations outside Africa.

### Functional mapping and annotation

Functional mapping and annotation (FUMA) [36] software was used to analyze GWAS data. FUMA, a web-based tool, works in two core processes: SNP2GENE and GENE2FUNC, using GWAS summary statistics as input. SNP2GENE identified functional SNPs and genes, outputting tables with significant SNPs, genomic risk loci, SNP positions, and mapped genes. It also generated Manhattan, Q-Q, and interactive regional plots, similar to those created in R-studio. GENE2FUNC provided outputs such as gene expression heat-maps, enrichment of differentially expressed gene sets in specific tissues, and links to external biological databases for further information on input genes.

## 3. Results

### 3.1 Participant demographics

The parent study [8], reported detailed participant demographics, indicating that the majority of respondents were female. The mean age of the GWAS participants (282 persons) was 35.1 years with a standard deviation (SD) of 9.3. A total of 178 participants were drawn from the rural Masaka TASO clinic and 104 participants were from the semi-urban Entebbe TASO clinics.

### 3.2 Quality control of raw reads

The quality of the raw data from the geno-typing array was assessed using GenomeStudio 2.0 software and is summarized in Table 1.

**Table 1:** quality metrics used for cleaning raw reads after genotyping from 282 samples.

| Quality metric |  |  |
| --- | --- | --- |
| Sample call rate ($> 0.95$) | Number of passed samples | 278 |
|  | Number of failed samples | 4 |
|  | Percentage success | 98.5% |
| GenTrain score ($0.0 - 1.0$) | Number of SNPs with score $\Rightarrow 0.5$ | 2,247,439 |
|  | Number of failed SNPs | 24,064 |
|  | Percentage success | 98.4% |

**Sample Call Rate:** The percentage of successful genotype calls for each sample, meaning 95% of calls per sample must be successful to pass quality control).
**GenTrain Score:** A score assessing the quality of SNP genotype clusters, where scores closer to 1.0 indicate better clustering and reliability of the SNP call. A score $\geq 0.5$ provides an optimal trade-off between sensitivity, including SNPs likely associated with the disease and specificity, avoiding false positives.

### 3.3 Quality control and data cleaning

The sequential steps undertaken to ensure high-quality genomic data are outlined in **Figure 2**, including pre-imputation and post-imputation QC procedures prior to GWAS analysis involving 282 participants and 2,247,439 SNPs. The final dataset comprised 275 participants (134 MDD cases, 141 controls) and 409,768 SNPs.

#### Principal components analysis of study cohort and its comparison with global reference populations from the 1000 genomes project

We conducted PCA to examine the genetic population structure within the study cohort. Figure S1.png in supplementary material is of the first two principal components (1 and 2) and it revealed a tightly clustered group of individuals, indicating a relatively homogeneous genetic background across the majority of participants. However, three individuals (samples 10, 62, and 5) were observed as clear outliers, positioned at a substantial distance from the main cluster. These were identified and removed from the dataset prior to GWAS analysis, as they could have reflected underlying population substructure due to ancestral admixture or differences in genetic ancestry.

Alternatively, they could have resulted from batch effects or genotyping artifacts. These findings are consistent with the broader PCA comparison including global reference populations, in which the study cohort aligned with African ancestry groups in the plot of principal components (PC) 1 against 2 as observed in **Figure 3**. We also plotted PC3 against PC4 which captured finer-scale substructure within Africa which had not been captured in PC1 and PC2, as observed in **Figure 3**. Together, these results support the genetic homogeneity of the study cohort.

### 3.5 Quantile-quantile plot visualization

The Q-Q plot in **Figure 4** illustrates the distribution of observed versus expected p-values under the null hypothesis of no association. The close alignment of the data points along the diagonal line indicates that the majority of the single nucleotide polymorphisms (SNPs) conform to the null expectation, suggesting no systematic inflation of test statistics.

**Figure 4:**
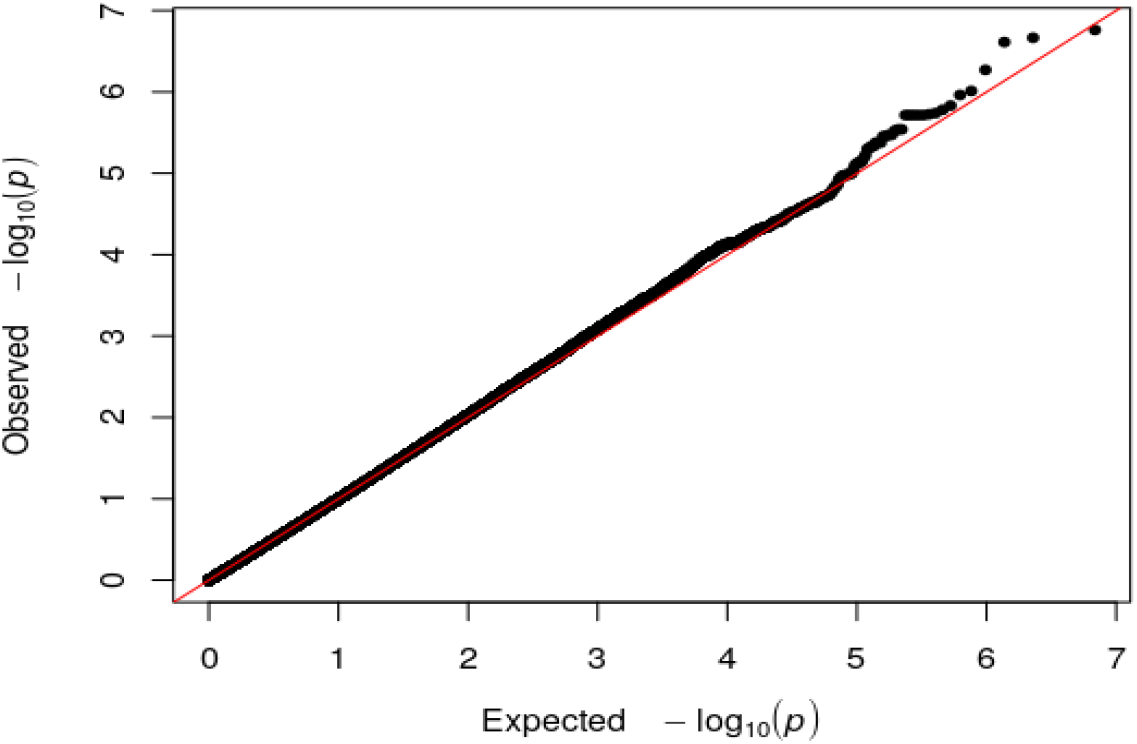
A Q-Q plot showing the extent to which the observed distribution of the p-values follows the expected p-values

Only a slight deviation is observed in the upper right tail of the distribution, where a small number of SNPs exhibit stronger-than-expected associations. This deviation suggests the presence of potential true signals of association, though weak and limited in number.

The genomic inflation factor (λGC = 0.9661), being close to 1, confirms the absence of substantial inflation or bias due to uncorrected population stratification, cryptic relatedness, or technical artifacts. Together, these results indicate that the statistical model used in the GWAS was well-calibrated and that any signals detected are unlikely to be driven by confounding structure in the data.

### 3.6 Genome-wide association and functional mapping and annotation results

We did not find any significant SNPs at the genome-wide significance threshold (p≤5×10^−8^). However, at a suggestive significance level of (p≤1×10^−6^), three SNPs were identified as potentially associated with MDD in this cohort. The Manhattan plot in **Figure 5** shows SNP association results at this threshold, with the most significant being rs4375916 on chromosome 2 (p=1.086×10^−7^). **Table 2** shows the leading 3 independent SNPs from the analysis that could be worth further investigating.

**Figure 5:**
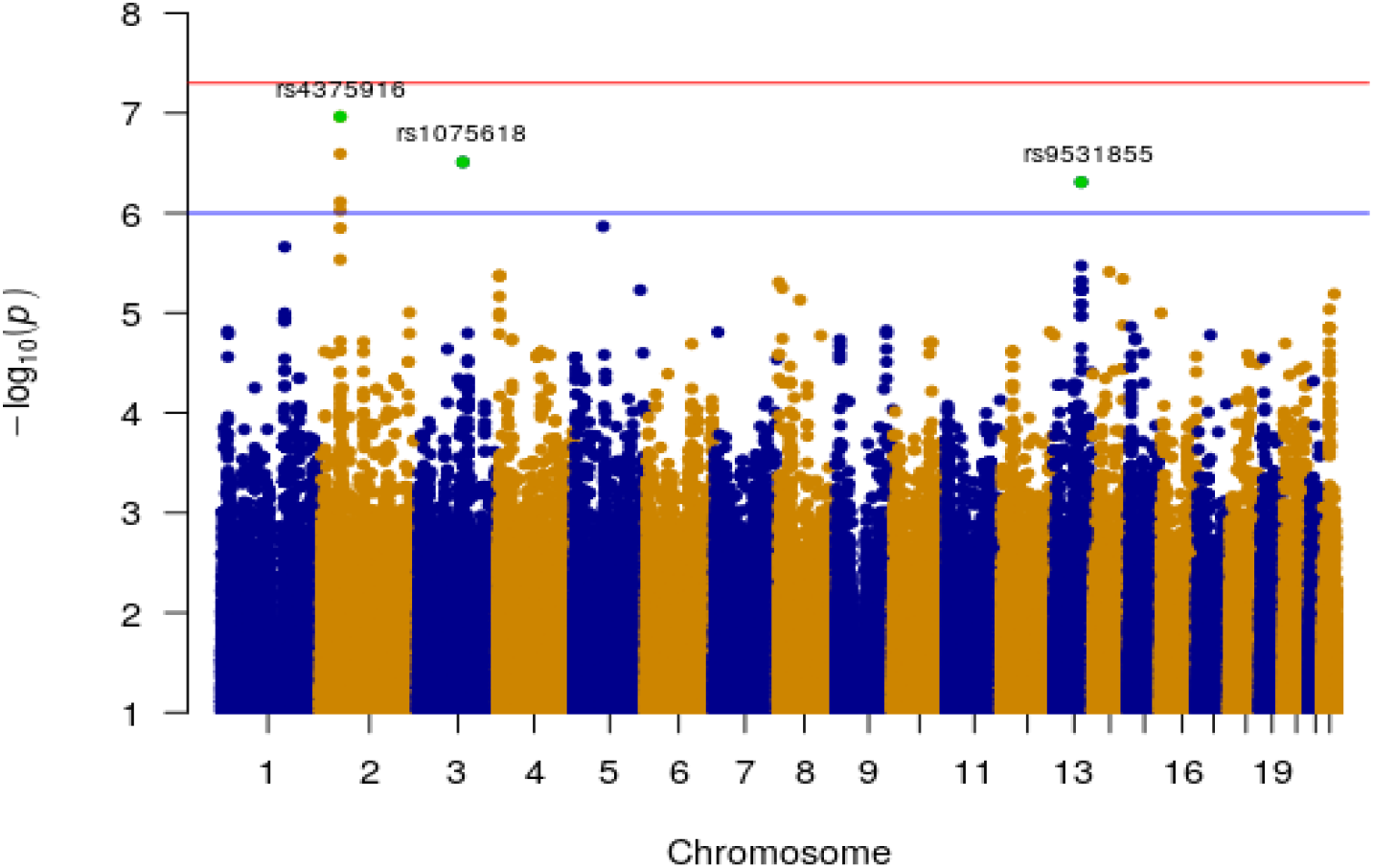
Manhattan plot displaying genome-wide significant SNPs across chromosome 1 to 22, at the suggestive threshold of (p≤1×10^−6^) as highlighted by the blue line. The red line is the known genome-wide significance threshold of (p≤5×10^−8^). The SNPs highlighted in green are the 3 independent leading SNPS

**Table 2.** A table showing the three leading SNPs from genome-wide analysis.

| rs ID | Chr | BP | Allele 1 | Allele 2 | MAF | Beta | S E | P-value | Region | Nearest gene |
| --- | --- | --- | --- | --- | --- | --- | --- | --- | --- | --- |
| rs4375916 | 2 | 48445680 | A | G | 0.466 | 0.223 | 0.041 | $1.086 \times 10^{-7}$ | ncRNA_intronic | *AC079A807.4* |
| rs1075618 | 3 | 111797917 | G | A | 0.304 | 0.255 | 0.049 | $3.093 \times 10^{-7}$ | intronic | *TMPRSS7* |
| rs9531855 | 13 | 86405753 | G | T | 0.240 | 0.271 | 0.053 | $4.906 \times 10^{-7}$ | intergenic | *SLITRK6* |

rs ID: Reference SNP identifier, Chr: chromosome, BP: base position, SE: standard error, MAF: minor allele frequency.

#### Functional mapping and annotation

Functional mapping and annotation identified three SNPs located within three genomic risk loci, based on a suggestive significance threshold (p ≤ 1 × 10^−6^). These loci, together with the 74 SNPs in linkage disequilibrium (LD) with the lead variants, were collectively mapped to 49 protein-coding genes using positional, eQTL, and chromatin interaction approaches. Of these, 28 genes demonstrated differential expression across 54 tissue types, as shown in Supplementary Figure S2. This heat-map presents gene expression levels across tissue, with each row representing a gene and each column a tissue type; blue indicates low or expression, while red indicates high expression. Among the 28 differentially expressed genes, *SLITRK6* and *RN7SKP224-FOXN2* showed suggestive genome-wide significance, as they were linked to two of the suggestively significant SNPS. Additional supplementary data have been included to support and expand on these findings. These include *Genomic risk loci*.*xlsx*, which summarizes the genomic risk loci identified based on independent lead SNPs and linkage disequilibrium (LD) structure; *linkage disequillibrium*.*xlsx*, which contains LD information on significant SNPs and SNPs in LD with the significant SNPs; and *genes*.*xlsx*, which summarises the list of genes mapped using FUMA’s positional, expression Quantitative Trait Locus, and chromatin interaction strategies. Furthermore, *gwascatalog*.*xlsx* presents the GWAS catalog gene enrichment results which summarizes the studies in which associated findings have been found.

#### Enrichment of input genes in gene sets

Examination of the enrichment input gene sets revealed the genes to be involved in a number of activities including regulation of pre-synapse organization, positive regulation of synapse activity, and maintenance of DNA repeat elements. This implies that genes associated with MDD are more frequently involved in synaptic function and genome stability than would be expected by chance, highlighting key biological mechanisms that may contribute to MDD warranting further investigation. These findings are illustrated in the supplementary figures provided in *positional_gene_sets_FUMA*.*docx* and are further summarized in *Gene set enrichment analysis*.*xlsx*, which details the significantly enriched biological pathways and processes, the overlapping genes, their statistical significance, and links to the corresponding Molecular Signatures Database (MSigDB) gene sets.

### 4.0 Discussion

This preliminary study explored the genetic risk of MDD in 282 PLWHIV attending ART clinics at TASO Entebbe and TASO Masaka in Uganda. With a modest sample size, the GWAS analysis identified 3 suggestively significant SNPs in 3 genomic risk loci associated with MDD. These SNPs together with the 74 other SNPs in LD with the 2 SNPs mapped to 49 protein-coding genes, with 28 expressed in 54 tissue types, including the brain.

The low number of risk loci, compared to larger studies in European ancestry populations, is likely due to the small sample size (275 participants after quality control). The study had limited statistical power, with only a 5.7% chance of detecting a true effect [37]. Despite the study’s low statistical power, three significant SNPs were identified using a relaxed significance threshold of (p≤ 1×10^-6^), which is less stringent than the commonly used threshold of (p≤8×10^-5^) for GWAS studies [32]. This adjusted threshold is often applied in underpowered studies and exploratory studies [38] to generate hypotheses that will be tested in larger studies [39], minimizing false negatives while ensuring that potential findings are not missed [40].

Worthy of note, among the three suggestively significant SNP, while none has been previously identified as associated with MDD, they are in linkage disequilibrium (LD) with SNPs previously reported in GWAS studies of other psychiatric conditions of Alzheimer’s disease and alcohol dependency–conditions that are comorbid with MDD [41], [42]. For instance, rs4375916 is in LD with rs2221714, a SNP previously described to have a significant association with Alzheimer’s disease [41]. This LD suggests that rs4375916 may tag [43] a genetic signal shared across these conditions. Major depressive disorder and Alzheimer’s disease have been reported to have a shared polygenic component [44], [45], [46] and also share common neurobiological abnormalities [45], [46]. However some studies found no evidence to support a common polygenic structure for Alzheimer’s and MDD [47], which was in contrast with what some studies have reported [42], [48]. The inconsistency in these findings highlights the need for larger, more comprehensive studies to establish conclusive evidence regarding genetic associations with these disorders. Individuals of African American ancestry have been reported to be at higher risk of Alzheimer’s than persons of European ancestry for reasons that may include economic disparities, cardiovascular health, quality of education, and biases in the methods used to diagnose Alzheimer’s [48]. Such studies are essential as they could enable reliable precision medicine approaches in persons with considerable African ancestry. Even though some studies have reported shared etiology between MDD and Alzheimer’s, we cannot confidently conclude that Africans or African Americans could be at higher risk for MDD as in Alzheimer’s.

The rs4375916 mapped to the *RN7SKP224 - FOXN2* gene, a pseudogene, which has also been identified in Alzheimer’s disease [41], however its role in Alzheimer’s disease has not been established as yet and no literature has been found to define its role in MDD or the brain tissue and related organs. This complexity may be particularly important for PLWHIV, who are already at a higher risk for MDD and other psychiatric disorders due to HIV related neuroinflammation, ART-induced neurochemical changes, and psychosocial stressors [49], [50]. There is a need for replication of rs4375916 in larger, sufficiently powered studies in order to allow further investigation of its role in the development of MDD among PLWHIV. Plausibly, HIV related immune dysregulation and chronic inflammation may interact with genetic predispositions to influence neuropsychiatric outcomes [49]. While rs4375916 may not directly be linked to HIV disease progression, its role in modulating neural or inflammatory pathways may render its effects more pronounced in a pro-inflammatory state such as HIV infection, thereby increasing susceptibility to MDD in this population.

Another suggestively genome-wide significant SNP, rs9531855, is in LD with rs9547398, which has also been previously identified in a study that conducted a meta-analysis of genetic influences on initial alcohol sensitivity [51]. This LD suggests that rs9531855 may tag [43] a genetic signal shared across MDD and initial alcohol sensitivity. This is not surprising, as several studies have reported that depression and alcohol use disorders (AUD) often occur together [51], [52]. Research indicates that higher alcohol consumption, influenced by genetics and environmental factors, increases the risk of developing major depression [53]. Alcohol use disorder and major depression each double the risk of the other [53], [54], and numerous studies show a strong association between alcohol dependence and depression [55]. In PLWHIV, the coexistence of depression and alcohol use disorder (AUD) presents particularly serious challenges, as both conditions independently contribute to reduced adherence to ART, accelerated HIV disease progression, and diminished quality of life [56], [57]. Depression in the context of AUD may further exacerbate alcohol consumption, as individuals may use alcohol to self-medicate depressive symptoms, thereby creating a reinforcing cycle [53]. Although most existing literature on the genetics of MDD and AUD has not been HIV specific, the mechanisms are likely to be amplified in HIV populations, where psychosocial stressors, HIV related neuroinflammation, and ART related side effects already increase vulnerability to psychiatric disorders. There are two possible explanations for the association between alcohol use disorders and major depression; first, it may be that both disorders have common underlying genetic and environmental factors that jointly increase the risk of both disorders. Second, the two disorders may have a causal effect, with each disorder increasing the risk of developing the other [52]. Understanding how these pathways operate in PLWHIV is crucial, given the added burden of HIV-related biological and social stressors on mental health outcomes.

The rs9531855 mapped to the *SLITRK6* gene that possesses variants which have been previously identified to have a role in bipolar disorder [55]. Bipolar disorder and (MDD) can sometimes be mistaken for one another, even by mental health professionals, due to the presence of depressive episodes in both conditions [58], [59]. Bipolar disorder and MDD have been identified to have several overlaps especially in their genetic epidemiology and molecular genetics [41], [60], [61], [62]. This is particularly relevant in HIV settings, where psychiatric symptoms may be misattributed to HIV related neurocognitive disorders or psychosocial stressors. Therefore, disentangling genetic susceptibility from HIV related or treatment related psychiatric manifestations is a critical step toward precision medicine in HIV mental health.

The enrichment analysis of the input gene sets suggests that genes associated with MDD play important roles in synaptic function and genome stability. Specifically, the genes were found to be involved in processes such as the regulation of pre-synapse organization, enhancement of synapse activity, and maintenance of DNA repeat elements. These findings are consistent with previous research that links disruptions in synaptic signaling to the development of depression [63], [64]. Synapses are crucial for communication between brain cells, and their proper function is essential for mood regulation [65]. Additionally, the involvement of genes in maintaining genome stability suggests that DNA integrity may also play a role in MDD. Together, these results point to biological pathways that may underlie the disorder and could serve as potential targets for future research or therapeutic interventions. In the context of PLWHIV, these pathways may be particularly relevant since HIV infection and ART can impact neuronal function, neuroinflammation, and oxidative stress, potentially disrupting synaptic signaling and neurotransmission, which are critical for mood regulation [66]. Moreover, HIV is associated with increased genomic stress and impaired DNA repair, which may exacerbate vulnerabilities linked to the maintenance of genome stability observed in MDD-associated genes [67]

A limitation of this study was the modest sample size of 282 participants on account of funding constraints. This resulted in an underpowered study (study power - 5.7%) increasing the likelihood of false-positive results. Small sample size is a frequent problem in studies of polygenic diseases and can result in insufficient power to detect minor contributions of one or more alleles. Similarly, small sample sizes can provide imprecise or incorrect estimates of the magnitude of the observed effects. For a highly powered GWAS for polygenic diseases, over thousands and millions of study participants are needed to have a conclusive study with no false positives associated to the discoveries [63]. Lastly, confounding may lie in the fact that most participants had comorbid psychiatric diagnoses, as in the case in most samples of psychiatric disorders. However, data on psychiatric comorbidities were not collected. Overall, the research into the genetics of MDD is limited especially in native African ancestry. As such, this study provides potentially useful preliminary data and methodological approaches to inform future investigations of this important area despite being unable to detect any significant associations in any of our analyses at known thresholds. Findings also present plausible evidence that with a larger sample size, there is a potential to identify more independent risk loci and significant SNPs associated with MDD in an African ancestry population.

## 5. Conclusion

This study did not identify any genome wide significant SNPs or genetic risk loci, most likely due to a small sample size, and consequent lack of statistical power. However, by applying a relaxed genome-wide suggestive threshold of (p≤1×10^-6^), we identified 3 suggestively genome-wide significant SNPs in 3 genetic risk loci, and none of these have been previously found to be associated with MDD. Importantly, our study was conducted among PLWHIV, a population burdened by depression due to HIV related biological and psychosocial factors. By highlighting suggestive genetic signals in this group, we underscore the need to investigate how host genetics interact with HIV infection, immune activation, and treatment effects to shape psychiatric outcomes. While preliminary, our findings demonstrate the feasibility of conducting GWAS in PLWHIV and provide a foundation for larger studies across African populations. Future work in larger HIV cohorts will be critical for disentangling the contributions of genetic risk and HIV related biology to depression, with the potential to inform targeted mental health interventions and precision medicine approaches in HIV care.

## Data Availability

The dataset presented in this article are currently not publicly available as further analysis is ongoing to address additional research questions. The datasets can be made available from the corresponding author on reasonable request.

## Declarations

### Ethical approval and consent to participate

This study was approved by the School of Biomedical Sciences Research Ethics Committee, Makerere University (number: SBS-2021-48), the Science and Ethics Committee of the Uganda Virus Research Institute Research (UVRI-REC - #GC/127/20/12/800) and the Uganda National Council for Science and Technology (UNCST -#HS 1053). All participants provided written informed consent in the EDCTP for their samples to be used in future genetic research. This study did not have to seek consent to participate as a waiver of consent had been obtained from the EDCTP sub-genetics [25]. The research was conducted in accordance with the Declaration of Helsinki.

### Clinical trial number

Not applicable

### Consent for publication

Not applicable.

### Authors’ contributions

Conceptualization, O.N.T., AK., EK., SMJH., and SS; Analysis: methodology, ONT., AK., and SF; validation, ONT., RG., SMJH., MS., CN., Mark.K., Martin.K., EK., AK., SF., SS., and Elly.K.; writing—original draft preparation, ONT; funding acquisition, EK, SS, SMJH, Mark.K,EK., AK., ONT., All authors have read and agreed to the published version of the manuscript.

## Acknowledgments

We acknowledge BioTeam for providing computational resources that were instrumental in the analysis of the data presented in this study. Research reported in this publication was supported by the South African Medical Research Council (SAMRC) Genomics of Brain Disorders (GBD) Unit. The content hereof is the sole responsibility of the authors and does not necessarily represent the official views of the DSI/SAMRC. Gratitude to the African Centre of Excellence, Infectious Diseases Institute and the Department of Immunology and Molecular Biology, College of Health Science, Makerere University and the Brain Health Project D43NS118560, for training and mentorship received from them to enable complete this research.

